# Remote, self-administered, smartphone cognitive testing in a registry-based cohort: Feasibility, reliability, and validity findings

**DOI:** 10.1101/2025.10.28.25338686

**Authors:** Sreya Dhanam, Mark Sanderson-Cimino, Jack Carson Taylor, Emily W. Paolillo, Ray Fregly, Winnie Kwang, Paul Maruff, Amy Wise, Hilary W. Heuer, Leah K. Forsberg, Joel H. Kramer, Bradley F. Boeve, Howard J. Rosen, R Scott Mackin, Michael W. Weiner, Rachel L. Nosheny, Adam L. Boxer, Adam M. Staffaroni, the Brain Health Registry

## Abstract

**Background:** Remote, smartphone-based cognitive testing may improve access to cognitive assessments for Alzheimer’s disease and related dementias. We evaluated the feasibility, reliability, and validity of unsupervised smartphone-based cognitive tests in a registry-based cohort.

**Methods:** Adults without a record of cognitive impairment (N=1,815; ages 18-92) were recruited from the UCSF Brain Health Registry to complete unsupervised ALLFTD-mApp cognitive tasks three times over two weeks. Reliability was assessed with correlations between sessions. Linear regression models tested associations of ALLFTD-mApp tasks with demographics, self- and informant-rated cognitive concerns (Everyday Cognition Surveys; ECog), and web-based cognitive testing (CogState Brief Battery; CBB).

**Results:** Adherence was high (82.2%) and usability favorable. Test-retest reliability was moderate to strong (ρs = 0.61–0.85, all ps < .001). Lower ALLFTD-mApp scores were associated with older age, lower education, cognitive concerns, and worse CBB performance.

**Conclusion:** Findings support the feasibility, reliability, and validity of the ALLFTD-mApp in adults without a record of cognitive impairment.

## INTRODUCTION

The prevalence of neurodegenerative diseases is expected to increase 2.5-fold by 2050, with corresponding rises in medical and economic costs [1,2]. Early identification of cognitive changes can help mitigate these costs by allowing for prevention, intervention, and improved care [3]. In individuals that develop neurodegenerative disease, repeated cognitive evaluations are often needed to evaluate progression, or in the era of disease modifying therapies, to monitor the effect of therapies. These cognitive evaluations have traditionally required in-person visits, trained study staff, and a controlled environment, creating barriers to accessibility and scalability [4]. Digital technologies present an opportunity to collect cognitive data remotely, allowing for high frequency assessments, low participant and study staff burden, and greater real-world applicability [5,6].

Several smartphone- and tablet-based assessment platforms are being developed for remote assessment of cognition [7–10]. One such tool is the ALLFTD Mobile Application (ALLFTD-mApp), developed by ALLFTD academic investigators in partnership with Datacubed Health, using their Linkt Health platform. This tool includes a comprehensive battery of self-administered cognitive, motor, and speech assessments, the ability to capture passive data via smartphone sensors, and an infrastructure for survey deployment [11]. Previous studies have shown that the app demonstrates feasibility, reliability, and validity in natural history studies of frontotemporal dementia [11,12]. These studies, however, have primarily involved individuals already diagnosed with neurodegenerative disease and enrolled in ongoing research. It is important to evaluate how the app performs in adults without a record of cognitive impairment, and to examine how demographic factors such as age and education influence task performance. Establishing the usability and psychometric characteristics in this population, as well as association with other gold-standard cognitive tests, will inform construct validity and the potential for using digital health technologies for scalable remote data collection.

In the current study, we test the feasibility and usability of remotely deploying the ALLFTD-mApp to a large sample of adults, ages 18 to 90+, recruited through the UCSF Brain Health Registry (BHR) [13]. We examine test-retest reliability and construct validity of these smartphone cognitive tests by examining the relationship between ALLFTD-mApp scores and: 1) demographic characteristics of the sample; 2) performance on a separate, well-validated, web-based cognitive battery (Cogstate Brief Battery (CBB); [14,15]) which provides an independent measure of cognition against which to validate remote app performance; and 3) self- and informant-based subjective cognitive changes, which have been linked to future cognitive decline, and are increasingly recognized as indicators of early cognitive impairment [16–18]. The ALLFTD-mApp includes several cognitive tasks, each with its own score, and creating an aggregate measure can help simplify interpretation and use in subsequent analyses [19]. We therefore report on the development of an ALLFTD-mApp cognitive composite score that can be computed in future studies. To ensure the composite score is applicable across a range of ability levels, we incorporated data from a natural history study of FTD and applied item response theory to develop the score.

## METHODS

### Study Design and Participants

Participants were recruited from the UCSF Brain Health Registry, an internet-based registry platform designed to facilitate Alzheimer’s disease and aging research by collecting longitudinal data from volunteers [13]. The only inclusion criterion for BHR is being 18 years of age or older.

Eligibility for the ALLFTD-mApp study also required participants to self-report English as their primary language. Additionally, exclusion criteria for the current study included a self- or informant-reported diagnosis of mild cognitive impairment, Alzheimer’s disease, or dementia, as well as a history of taking AD-related medications, including donepezil (Aricept), rivastigmine (Exelon), memantine (Namenda), galantamine (Razadyne), or aducanumab (Aduhelm). Participants completed a measure of subjective cognitive complaints through the BHR; those scoring above established thresholds of subjective cognitive impairment (Everyday Cognition Scale; ECog >1.81 for self-report and ≥1.88 for informant-report) were also excluded. A total of 33,470 participants met these criteria and were informed about the mobile app study, and 1,829 completed consent and were enrolled in the study. Demographic data was available for 1,815 of these enrolled participants. Participants were expected to use their own smartphones (iOS or Android). The ALLFTD-mApp supports Android 6.0 and later (Marshmallow onwards) and is built for backward compatibility across device generations. The current iPhone application requires an iPhone 8 or newer (iOS 16.0 or later), although earlier releases supported older models.

To support the development of a cognitive composite score, 490 additional participants from the ARTFL LEFFTDS Longitudinal Frontotemporal Lobar Degeneration (FTLD) consortium (ALLFTD) who completed the same battery of smartphone cognitive tasks were included in composite score development. The subsample of ALLFTD participants with smartphone data has been described previously [11,12], and includes participants with familial and sporadic FTLD spectrum disorders. They present with a heterogenous set of symptoms, suspected underlying pathology, and clinical severity [20]. Developing the ALLFTD-mApp composite score using item response theory in a combined sample of ALLFTD and BHR participants ensures that the score represents a broad range of the underlying latent cognitive ability (see Supplementary Materials for details). In this study, estimates of feasibility, reliability, or validity were based solely on the BHR sample.

The study was approved by the UCSF Institutional Review Board and is conducted in accordance with the latest Declaration of Helsinki, including written informed consent from all participants.

### Procedure

The referral and enrollment process was conducted in a completely remote and unsupervised fashion. Referral waves from the BHR were spaced out approximately every two weeks from 2022 - 2024. Eligible BHR participants at each referral wave were first contacted by email. Those who did not respond were sent up to two reminder emails, 3 and 6 days after the initial contact. Interested participants were asked to complete an initial data sharing consent through the BHR portal. After consenting, they were provided with training materials, including instructional videos and documents to support app downloading and navigation. Next, participants were prompted to create personalized login credentials linking mobile app data to BHR data. Using these credentials, participants accessed the ALLFTD-mApp (compatible with both iOS and Android; https://www.datacubed.com). Informed consent of study procedures was then conducted through the app.

### ALLFTD Mobile App (Linkt Health) Assessment

Participants independently downloaded the ALLFTD-mApp onto their personal smartphone. The ALLFTD-mApp includes a multi-domain digital testing battery that was built on Datacubed Health’s Linkt Health app. The battery included self-administered tasks of cognition, speech and language, motor functioning, and questionnaires. The cognitive task and questionnaire data were the focus of this study. Similar to the protocol used in the ALLFTD study [12], participants were asked to complete three testing sessions over an 11-day period. Specifically, the first battery of tests was available for three days. This session was followed by a washout day (Day 4) during which no tasks were available. On the fifth day, the tasks became available again for three days, followed by another washout day (Day 8). On the next day, participants were asked to complete the battery for a third time, again with three days to complete the tasks. Each battery took approximately 30-45 minutes to complete. Participants were compensated with a $10 Amazon gift card upon completion of each session (up to $30 total). All participants were asked to enable banner notifications about upcoming testing sessions. Participants could also opt in to receiving additional text message reminders (n = 1718).

*Executive Functioning Tasks:* This study focuses specifically on the gamified versions of four cognitive tasks adapted from standard neuropsychological assessments, described in detail elsewhere [11]. Measures included Color Clash (Stroop), a task of cognitive inhibition based on the Stroop paradigm [21], Ducks in a Pond (Flanker), a task of attentional control adapted from the Eriksen Flanker Task [22]; Animal Parade (N-back), a task of working memory that builds on the n-back paradigm [23], and Card Shuffle (Card Sort), a task of cognitive flexibility modeled after the Wisconsin Card Sorting Test [24]. Each task was administered three times over the study period, and participants completed a brief practice trial for all tasks other than Card Sort. The Card Sort task, which was only administered once given its well-documented practice effects [25].

*Episodic Memory Task*: Humi’s Bistro, an associative memory task, requires participants to remember and match food and table order pairings. The task is adaptive, with task difficulty increasing or decreasing based on participant performance. The primary outcome is the mean number of correct responses across trials.

*Smartphone Composite Score*: The Episodic Memory Task, N-back, Flanker, and Stroop were combined into a smartphone composite score via item response theory using previously established methods [6] that are fully described in the Supplementary Materials. Card Shuffle was excluded because it was only administered once.

*12-item Everyday Cognition Survey:* On the app, participants completed a self-report measure of subjective cognitive concerns, the ECog-12 [26]. The ECog-12 was derived from the original 39- item version (ECog-39), which was developed and validated to assess early functional changes associated with cognitive decline [27]. Participants rate their perceived change in functioning across cognitive domains compared to 10 years ago on a 4-point scale from 1 (better or no change compared) to 4 (consistently much worse). The primary ECog-12 score is an average of their responses to each item. In addition, average scores for the memory and executive functioning subdomains were also calculated.

*Feasibility and User Experience Questionnaire:* Participants completed a questionnaire assessing user experience. This questionnaire was modeled after similar feasibility and usability surveys [8,28]. Participants rated their agreement with various statements on a 5-point Likert scale (1 = strongly disagree, 5 = strongly agree), including:

1. I trust that the study smartphone app keeps my personal data private and secure.
2. Overall, the task instructions were clear.
3. The app’s text is usually large enough to read comfortably.
4. I prefer to take cognitive tests using a smartphone rather than using pen and paper.

Additionally, participants provided feedback on task difficulty and time commitment through rating-scale questions. At the end of each session, participants reported any distractions during testing.

### BHR Web-based Assessments

*Cogstate Brief Battery:* As part of BHR study procedures, a subset of 560 participants also completed CBB tasks within a year of the ALLFTD-mApp study start date. The CBB is a computerized assessment that includes validated cognitive measures such as: the Detection Task (DET) which evaluates simple reaction time; the Identification Task (IDN) which evaluates choice reaction time; the One Card Learning Task (OCL) which evaluates visual learning and memory; and the One Back Task (ONB) which evaluates working memory [15]. These tasks have been shown to be reliable and sensitive to cognitive changes associated with neurodegenerative diseases [15,29]. They have also been shown to be responsive to general age-related cognitive changes [30]. Performance metrics included reaction time for DET, IDN, OCL and ONB tasks.

### Study Partner Assessments

*ECog-41:* Participants were asked to identify a study partner that interacts with them daily to give insight into their everyday cognition. Study partners (informants) were asked to complete a REDCap version of the ECog-41, a 41-item survey designed adapted from the ECog-39 [31]. The primary ECog-41 score is an average of all item responses (possible range per item = 1-4).

### Statistical Analysis

Data analyses were conducted using R, versions 4.3.1 and 4.5.0 with statistical significance set at p < .05. All cross-sectional analyses were based on the first session completed by each participant.

#### Associations with demographic characteristics

First, to understand the demographic and clinical characteristics of the study sample as well as to appreciate the psychometric characteristics of the performance measures from the tests contained in the ALLFTD app, descriptive statistics were used to summarize demographic and cognitive performance data.

Second, to examine relationships between the sample characteristics and performance on the ALLFTD-mApp, we fit linear regression models to assess the association of app-based task performance with demographics, including age, gender, and education. For visualization, linear models were used to plot relationships between age and task performance for most cognitive tasks. For the Card Sort task, both quadratic and generalized additive models were fit to examine the relationship with age. A generalized additive model provided a better fit based on visual inspection and was supported by a statistically significant nonlinear effect **(Figure 1).**

**Figure 1:**
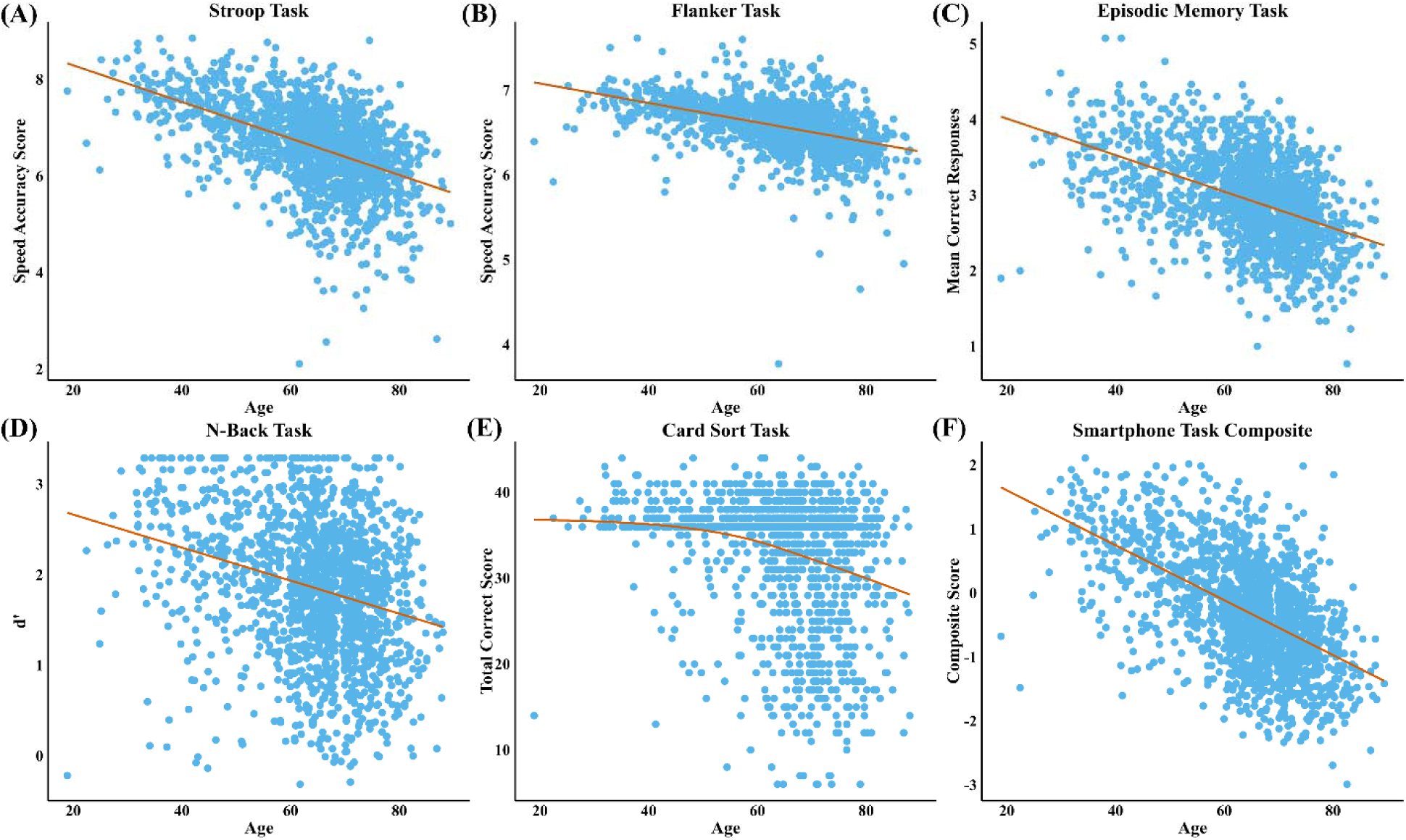
Associations between age and ALLFTD-mApp task performance. This figure displays scatterplots illustrating the relationships between participant age and performance on each of the five cognitive tasks included in the ALLFTD-mApp, as well as the overall smartphone task composite score. Linear regression lines are shown for most tasks; as a statistically significant nonlinear relationship was detected in the Card Sort Task (E), a generalized additive model fit is shown instead. Participants aged 90 or older were included in analyses but are not shown in this figure to reduce re-identification risk.

#### Associations with independent cognitive tests, and self- and informant-based reports of cognitive complaints

We then evaluated convergent validity by testing correlations between app-based task performance, ECog scores, and CBB measures. Due to non-normal distributions of some cognitive assessment data, Spearman’s rank correlation coefficients were fit to assess relationships between app-based tasks, ECog scores and CBB assessments. For analyses involving the CBB, we conducted a sensitivity analysis that included the time interval between the app and CBB assessments as a covariate. In a second sensitivity analysis, device type (iOS vs Android) was included as a covariate within regression models. We did not include demographic characteristics as covariates since our objective was to evaluate associations between app-based measures and criterion standards, rather than their incremental predictive value.

#### Test-Retest Reliability of App-Based Cognitive Tasks

Test-retest reliability of the ALLFTD-mApp tasks was assessed using spearman correlations. Correlations were calculated between timepoints (Sessions 1-2 and 2-3) for each task separately, using only participants who completed that task at all three timepoints (n = 1207-1252, depending on the task).

#### Adherence and Feasibility

Finally, we examined feasibility by calculating adherence rates and testing the impact of text reminders. Mean adherence rates and standard deviations were computed across all three sessions and by individual session. Adherence was calculated as the proportion of tasks that were completed. To evaluate the impact of text message reminders on adherence, a linear regression model was used to compare participants who received text message reminders to those who did not.

## RESULTS

### Participant Characteristics

Demographic characteristics of the sample (n = 1,815) are reported in **Table 1**. Participants had a mean age of 64 years (SD = 12; range 18-92). The mean years of education was 16 (SD = 2.3; range 6-20). 67% of participants identified as female, 33% as male, and 0.3% as another gender, consistent with overall BHR demographics [13]. The racial composition was predominantly White (83%) and not Latino (93%). A total of 314 study partners completed the ECog-41 to assess the participant’s cognitive function (see Supplementary Materials for demographic details).

**Table 1.** Demographic and cognitive characteristics by age group. This table presents demographic, device type, and cognitive characteristics for the full sample (N = 1,815), stratified by age group. **Abbreviations:** BHR, Brain Health Registry; ECog, Everyday Cognition questionnaire; CBB, Cogstate Brief Battery; DET, Detection task; IDN, Identification task; ONB, One-back task; OCL, One-card learning task; RT, reaction time; SD, standard deviation; NA, not available.

|  |  | Age Bin |  |  |  |  |  |  |
| --- | --- | --- | --- | --- | --- | --- | --- | --- |
|  | Overall | 18-29 | 30-39 | 40-49 | 50-59 | 60-69 | 70-79 | 80+ |
| **n** | **1815** | **10** | **68** | **149** | **238** | **698** | **548** | **104** |
| **Age (mean, SD)** | 64.21<br>(11.56) | 25.60<br>(3.06) | 35.38<br>(2.32) | 44.91<br>(2.81) | 55.11<br>(2.70) | 64.74<br>(2.62) | 73.50<br>(2.61) | 82.65<br>(2.88) |
| **Years Education** | 16.24<br>(2.35) | 15.00<br>(2.49) | 17.21<br>(1.69) | 16.98<br>(1.73) | 16.55<br>(2.18) | 15.90<br>(2.36) | 16.13<br>(2.48) | 16.77<br>(2.52) |
| **Gender (%)** |  |  |  |  |  |  |  |  |
| *Male* | 599<br>(33.0) | 5<br>(50.0) | 10<br>(14.7) | 32<br>(21.5) | 64<br>(26.9) | 220<br>(31.5) | 226<br>(41.2) | 42<br>(40.4) |
| *Female* | 1210<br>(66.7) | 5<br>(50.0) | 57 (83.8) | 115<br>(77.2) | 172<br>(72.3) | 477<br>(68.3) | 322<br>(58.8) | 62<br>(59.6) |
| *Other* | 6<br>(0.3) | 0<br>(0.0) | 1<br>(1.5) | 2<br>(1.3) | 2<br>(0.8) | 1<br>(0.1) | 0<br>(0.0) | 0<br>(0.0) |
| **Race (%)** |  |  |  |  |  |  |  |  |
| *Caucasian* | 1512<br>(83.3) | 5<br>(50.0) | 54<br>(79.4) | 116<br>(77.9) | 179<br>(75.2) | 584<br>(83.7) | 475<br>(86.7) | 99<br>(95.2) |
| *Multiracial* | 184<br>(10.1) | 1<br>(10.0) | 10<br>(14.7) | 21<br>(14.1) | 38<br>(16.0) | 73<br>(10.5) | 40<br>(7.3) | 1<br>(1.0) |
| *African American* | 43<br>(2.4) | 1<br>(10.0) | 2<br>(2.9) | 3<br>(2.0) | 4<br>(1.7) | 16<br>(2.3) | 15<br>(2.7) | 2<br>(1.9) |
| *Asian* | 56<br>(3.1) | 2<br>(20.0) | 2<br>(2.9) | 8<br>(5.4) | 13<br>(5.5) | 15<br>(2.1) | 15<br>(2.7) | 1<br>(1.0) |
| *Native American* | 5<br>(0.3) | 0<br>(0.0) | 0<br>(0.0) | 0<br>(0.0) | 1<br>(0.4) | 2<br>(0.3) | 2<br>(0.4) | 0<br>(0.0) |
| *Pacific Islander* | 1<br>(0.1) | 0<br>(0.0) | 0<br>(0.0) | 0<br>(0.0) | 1<br>(0.4) | 0<br>(0.0) | 0<br>(0.0) | 0<br>(0.0) |
| *Other* | 10<br>(0.6) | 0<br>(0.0) | 0<br>(0.0) | 1<br>(0.7) | 1<br>(0.4) | 7 (1.0) | 1<br>(0.2) | 0<br>(0.0) |
| *Latino* | 4<br>(0.2) | 1<br>(10.0) | 0<br>(0.0) | 0<br>(0.0) | 1 (0.4) | 1 (0.1) | 0<br>(0.0) | 1 (1.0) |
| **Ethnicity (%)** |  |  |  |  |  |  |  |  |
| *Latino* | 122<br>(6.7) | 1<br>(10.0) | 7<br>(10.3) | 18<br>(12.1) | 28<br>(11.8) | 43<br>(6.2) | 24<br>(4.4) | 1<br>(1.0) |
| *Not Latino* | 1690<br>(93.2) | 9<br>(90.0) | 61<br>(89.7) | 131<br>(87.9) | 210<br>(88.2) | 655<br>(93.8) | 522<br>(95.4) | 102<br>(98.1) |
| *Declined to State* | 2<br>(0.1) | 0<br>(0.0) | 0<br>(0.0) | 0<br>(0.0) | 0<br>(0.0) | 0<br>(0.0) | 1<br>(0.2) | 1<br>(1.0) |
| **Device Type (%)** |  |  |  |  |  |  |  |  |
| *iOS* | 1241<br>(68.7) | 7<br>(70.0) | 43<br>(57.3) | 92<br>(62.2) | 183<br>(72.6) | 467<br>(65.8) | 382<br>(72.9) | 67<br>(77.0) |
| *Android* | 565<br>(31.3) | 3<br>(30.0) | 32<br>(42.7) | 56<br>(37.8) | 69<br>(27.4) | 243<br>(34.2) | 142<br>(27.1) | 20<br>(23.0) |
| **Cognition** |  |  |  |  |  |  |  |  |
| **ECog Scores** |  |  |  |  |  |  |  |  |
| *BHR Web-Based ECog-12* | 1.29<br>(0.09) | 1.23<br>(0.16) | 1.22<br>(0.05) | 1.29<br>(0.12) | 1.30<br>(0.09) | 1.27<br>(0.05) | 1.30<br>(0.03) | 1.39<br>(0.26) |
| *ALLFTD App-Based ECog-12* | 1.28<br>(0.07) | 1.21<br>(0.23) | 1.21<br>(0.08) | 1.29<br>(0.11) | 1.31<br>(0.09) | 1.26<br>(0.02) | 1.28<br>(0.03) | 1.33<br>(0.19) |
| *Study Partner Ecog-41 (n=314)* | 1.30<br>(0.30) | NA | 1.08<br>(0.02) | 1.19<br>(0.04) | 1.16<br>(0.04) | 1.38<br>(0.45) | 1.32<br>(0.08) | 1.23<br>(0.10) |
| **App Executive Functioning Task Overall Score (n=1811)** |  |  |  |  |  |  |  |  |
| *Composite Score* | 0.08<br>(0.47) | 0.83<br>(0.72) | 1.19<br>(0.19) | 0.82<br>(0.20) | 0.51<br>(0.26) | 0.04<br>(0.12) | -0.27<br>(0.11) | -0.75<br>(0.18) |
| **CBB Scores (n=560)** |  |  |  |  |  |  |  |  |
| *RT DET* | 2.53<br>(0.03) | 2.52<br>(0.01) | 2.52<br>(0.06) | 2.51<br>(0.03) | 2.51<br>(0.03) | 2.54<br>(0.01) | 2.54<br>(0.03) | 2.57<br>(0.00) |
| *RT IDN* | 2.70<br>(0.02) | 2.71<br>(0.00) | 2.68<br>(0.04) | 2.68<br>(0.01) | 2.68<br>(0.02) | 2.70<br>(0.01) | 2.71<br>(0.02) | 2.74<br>(0.00) |
| *RT ONB* | 2.84<br>(0.03) | 2.73<br>(0.00) | 2.79<br>(0.05) | 2.82<br>(0.03) | 2.82<br>(0.04) | 2.85<br>(0.02) | 2.85<br>(0.02) | 2.90<br>(0.00) |
| *RT OCL* | 2.98<br>(0.03) | 2.52<br>(0.00) | 2.92<br>(0.03) | 2.96<br>(0.02) | 2.97<br>(0.02) | 2.98<br>(0.01) | 2.99<br>(0.02) | 3.02<br>(0.01) |

### Associations with demographic characteristics

Older age was significantly associated with poorer performance across all app-based tasks, except for Card Sort, which displayed a more variable, non-linear pattern across the lifespan (**Figure 1**). Specifically, older age was linked to worse performance on Stroop (β = -0.51, 95% CI [-0.51, -0.50]), Episodic Memory (β = -0.47, 95% CI [-0.47, -0.47]), Flanker (β = -0.44, 95% CI [-0.45, -0.44]), N-back (β = -0.26, 95% CI [-0.26, -0.26]), and Card Sort (β = -0.22, 95% CI [-0.25, -0.19]), all p < .001 (**Figures 1 and 2**). Higher education was associated with better performance on all tasks: Stroop (β = 0.08, 95% CI [0.06, 0.10]), Episodic Memory (β = 0.07, 95% CI [0.06, 0.08]), Flanker (β = 0.12, 95% CI [0.12, 0.13]), N-back (β = 0.12, 95% CI [0.11, 0.14]), and Card Sort (β = 0.16, 95% CI [0.01, 0.31]), all p < .001. The overall smartphone task composite score was significantly associated with older age (β = -0.55, 95% CI [-0.55, -0.55], p < .001) and higher education (β = 0.12, 95% CI [0.11, 0.14], p < .001). There were no significant associations with gender and the overall composite or any individual task (β’s < 0.03, p’s > .28).

**Figure 2:**
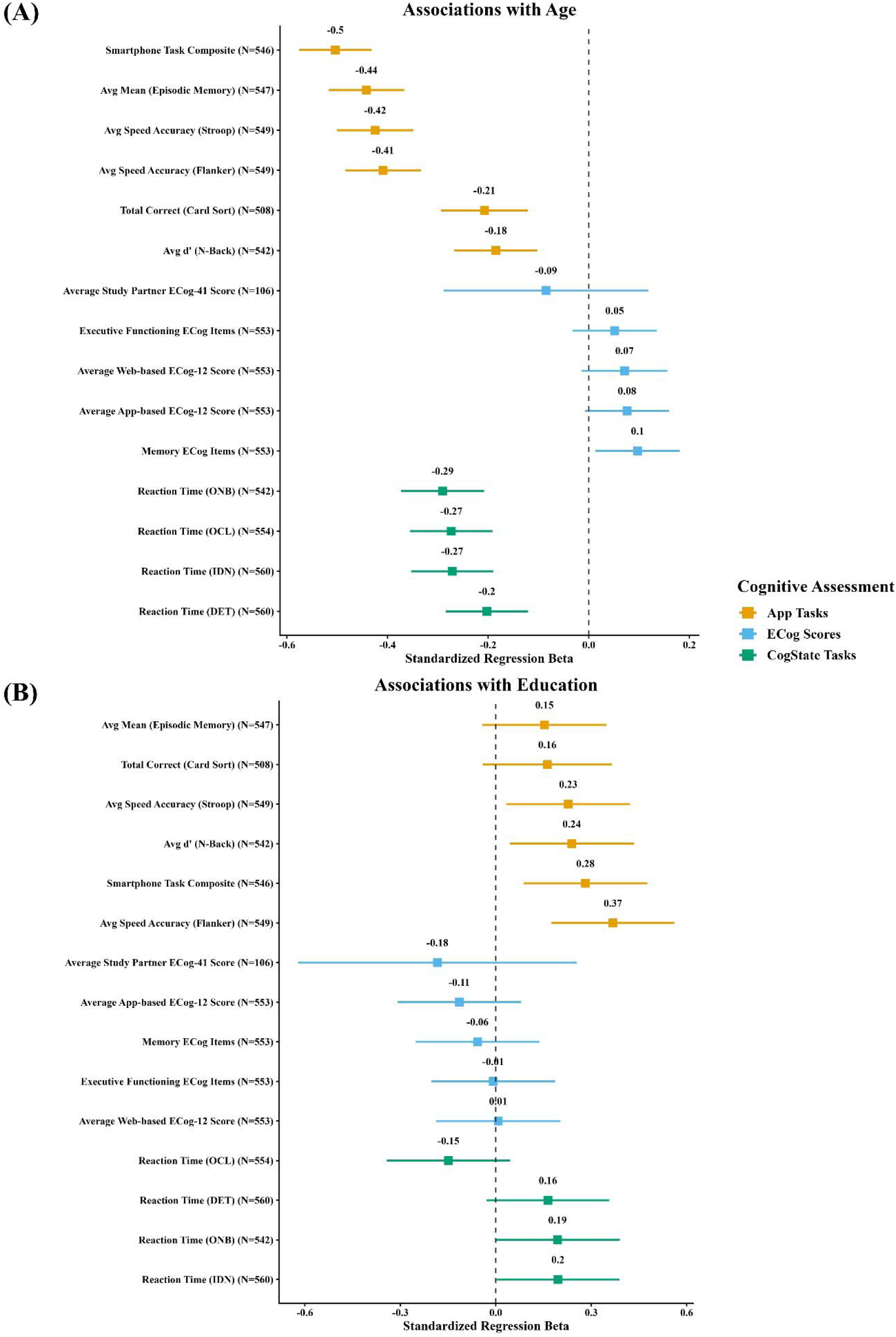
Associations of age and education with digital cognitive tests and subjective cognitive complaints. The two panels display standardized regression coefficients (β) and 95% confidence intervals for the associations between cognitive outcomes and (A) age and (B) education. Associations are adjusted for education and gender. Participants included in these analyses completed both the smartphone and CBB tasks.

### Associations with independent cognitive tests

ALLFTD-mApp tasks of processing speed and executive functioning showed associations with CBB tasks of the same domain in the expected directions (**Figure 3**). The ALLFTD-mApp composite score was strongly associated with CBB metrics of speed and executive functioning (e.g., ONB reaction time; ρ = -0.47, 95% CI [-0.54, -0.41], p < .001). The ALLFTD-mApp Episodic Memory task was associated with the CBB memory task (OCL; ρ = -0.15, 95% CI [- 0.23, -0.06], p < .001) as well as with other CBB tasks (ρ*’s* = -0.27 to -0.30, p’s < .001).

**Figure 3:**
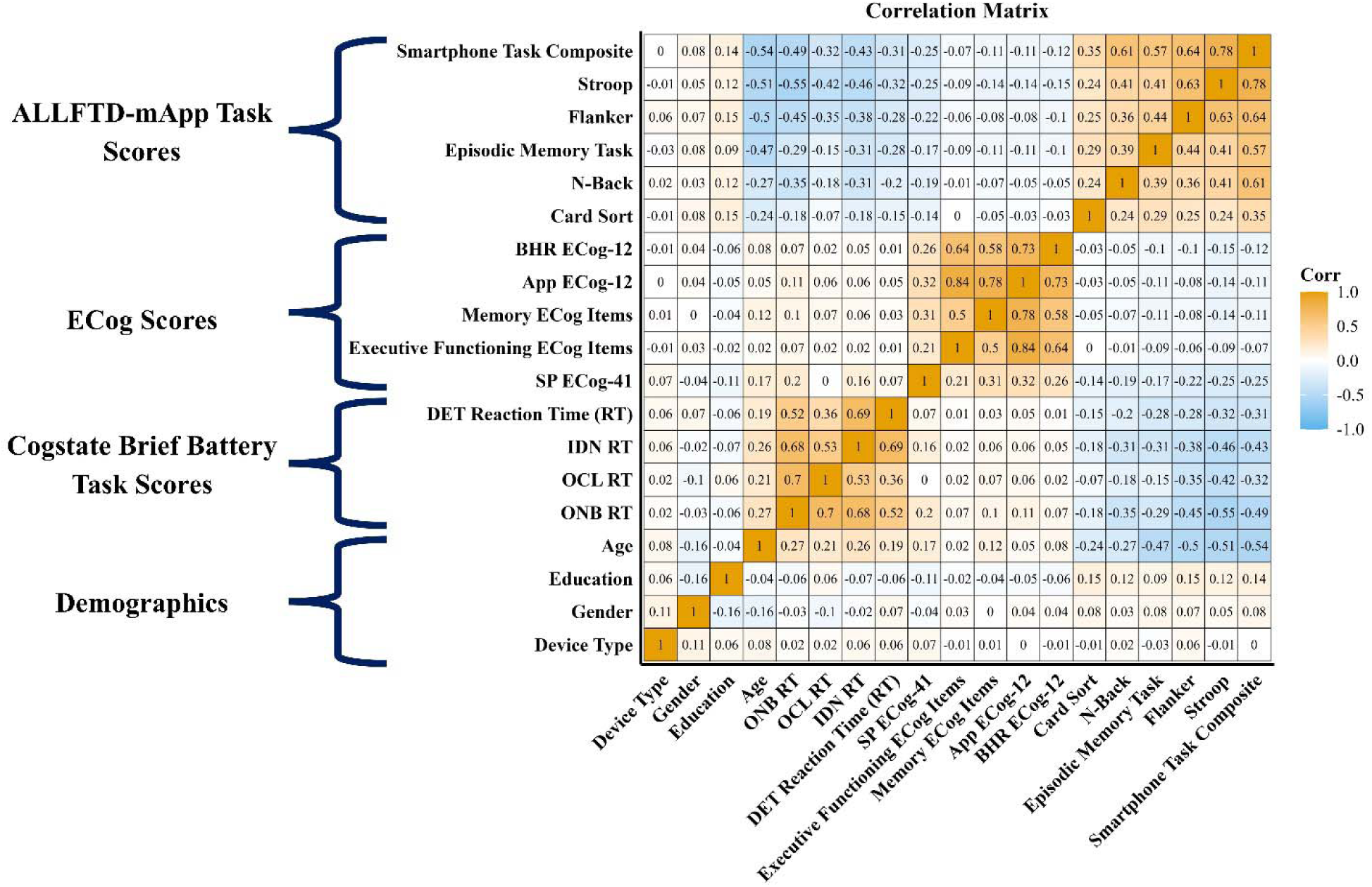
Correlation matrix of ALLFTD-mApp task performance, ECog scores, CBB scores, and demographics. This figure displays Spearman correlation coefficients among demographic variables, ALLFTD- mApp cognitive task scores, participant and study partner ECog scores, and CBB task performance. Variables are grouped into four domains: ALLFTD-mApp task scores, ECog scores, CBB task scores, and demographics. Warmer colors (orange) indicate stronger positive correlations; cooler colors (blue) represent stronger negative correlations. As expected, age showed moderate negative correlations with most app-based task scores. Education was positively associated with both app and web-based task performance. Strong internal consistency was observed within the ECog domain and among the app-based executive functioning tasks. The smartphone task composite showed moderate correlations with both web-based ECog scores and CBB metrics, supporting the validity of the app-based assessments.

### Associations with self- and informant-based reports of cognitive complaints

ECog scores derived from the app-based administration and the BHR web-based administration were highly correlated even after adjusting for the time between administration of these measures (ρ = 0.73, 95% CI [0.70, 0.75], p < 0.001; **Figure 3**). Moderate, statistically significant associations were observed between app-administered ECog and app-administered informant ECog scores (ρ = 0.31, 95% CI [0.20, 0.41], p < 0.001).

Greater self-reported subjective cognitive complaints (ECog-12) were associated with worse performance on the Stroop, Flanker, and Episodic Memory tasks (ρ*’s* ≥ -0.08; p’s < .05). Worse self-reported memory functioning was associated with poorer performance on the Episodic Memory task (ρ = -0.11, 95% CI [-0.16, -0.11], p < .001) as well as worse performance on executive functioning tasks (ρ*’s* = -0.05 to -0.14, p’s ≤ .05). The executive functioning subscale was negatively correlated with performance on Flanker (ρ = -0.06, 95% CI [-0.11, - 0.01], p < .05) and Stroop (ρ = -0.08, 95% CI [-0.13, -0.04], p < .001).

Informant concerns about participants’ cognitive abilities were also associated with worse performance on app-based tasks (**Figure 3**). In this subsample, the magnitude of associations was similar to those observed for participant self-report: Stroop (ρ = -0.24, 95% CI [-0.36, - 0.13], p < .001), Flanker (ρ = -0.20, 95% CI [-0.31, -0.09], p < .001), Episodic Memory (ρ = -0.16, 95% CI [-0.27, -0.03], p = .006), N-back (ρ = -0.16, 95% CI [-0.27, -0.05], p = .009), Card Sort (ρ = -0.13, 95% CI [-0.24, -0.01], p = .03), and the overall composite score (ρ = -0.23, 95% CI [-0.33, -0.12], p < .001).

### Operating System and ALLFTD App Task Performance

The Apple (iOS) operating system was associated with better performance on a combined speed/accuracy outcome from the Flanker task, as compared to the Android operating system (β = 0.10, 95% CI [0.07, 0.12], p < .001). There were no other statistically significant differences in performance by operating system (p’s > .05). In sensitivity analyses, Flanker associations with demographics, CBB, and the ECog-12 tasks remained similar after adjusting for operating system.

### Test-Retest Reliability of App-Based Cognitive Tasks

The Composite Score (ρ = 0.80 for Sessions 1-2; ρ = 0.85 for Sessions 2-3), Stroop (ρ = 0.78- 0.85), and Flanker (ρ = 0.82-0.84) tasks showed strong reliability, while the Episodic Memory (ρ = 0.67-0.71) and N-back (ρ = 0.67-0.72) tasks demonstrated moderate reliability. All correlations were statistically significant (p < .001), with 95% confidence intervals indicating robust reliability estimates across tasks (see Supplemental Materials for full estimates).

### Adherence and Feasibility

Adherence was high, with 86% of participants completing at least one full session. Overall adherence was calculated as the number of completed tasks out of all possible tasks across all three sessions and averaged 82% (SD = 35%). Adherence declined slightly across sessions: 88% in Session 1 (SD = 25%), 81.9% in Session 2 (SD = 37%), and 77% in Session 3 (SD = 41%).

Participants who received any SMS reminder had significantly higher adherence than those who received none (β = 0.19, p < .001), representing a 19 percent increase.

Participants responded to six feasibility and user experience questions. Across age groups (<65, 65-75, and 75+), most participants reported that the app’s text was large enough to read comfortably (∼95%), instructions were clear (∼90%), and the 30-minute session length was acceptable (80%) (**Figure 4**). A majority (∼70%) agreed or strongly agreed they preferred smartphone-based testing over pen and paper and felt their data were private and secure. Most participants rated task difficulty as “Somewhat Difficult” to “Very Easy,” with similar patterns observed across age groups.

**Figure 4.**
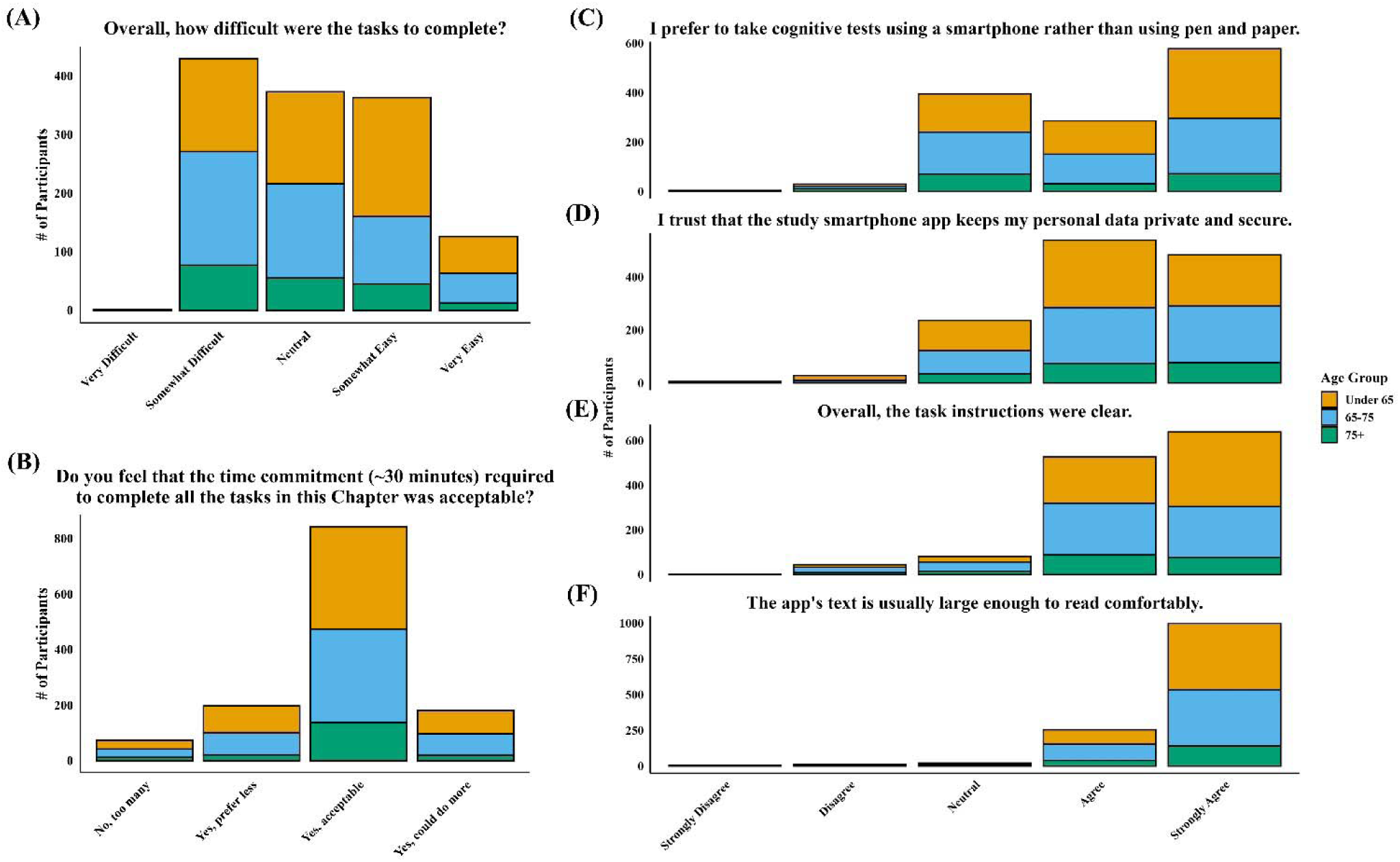
*Participant feedback on smartphone-based cognitive testing by age group.* Participant responses to six feasibility and usability questions, stratified by age group (<65, 65– 75, 75+). Across all age groups, most found task instructions clear (E), text readable (F) and session length acceptable (B). The majority of participants preferred smartphone-based testing over pen and paper (C) and felt their data were secure (D). Task difficulty was generally rated as “Somewhat Difficult” to “Very Easy,” with similar patterns across age groups (A).

A minority of participants reported distractions during testing: 28% in session 1, 21% in session 2, and 21% in session 3.

## DISCUSSION

In this study we provide evidence for feasibility and usability through high adherence and positive user feedback. Reliability was demonstrated in participants who completed the tasks up to three times over an 11-day period, with results showing moderate to strong associations between assessments. Construct validity of these mobile cognitive tasks was supported through associations of the ALLFTD-mApp tasks and demographics, self- and informant-rated cognitive concerns, and an independent web-based cognitive testing battery. These findings highlight the potential of digital tools to expand access to cognitive assessments.

This study supports the feasibility and scalability of remote smartphone-based cognitive testing. In prior ALLFTD-mApp studies, research coordinators assisted participants with app download and set up. Here we show that it is feasible for participants to download and navigate through the app without the support of study staff, although participants could contact the study team via email if assistance was required. This greatly increases the potential for expanding access to such a technology across larger healthcare systems. Despite the lack of contact with the study team, adherence was over 80%, which is comparable to ALLFTD-mApp adherence reported in a FTD natural history study [11] and other unsupervised remote cognitive testing protocols, including the BRANCH app, ARC app and NIH Mobile Toolbox [8,32,33]. These estimates are also consistent with rates described in a recent scoping review of similar studies (reported range: 74-93%) [34].

Usability of the technology was supported by user experience survey results, which indicated that most of the participants strongly preferred smartphone-based testing over pen-and-paper assessments. Other feasibility metrics were also encouraging, including participant reports that the instructions were clear and the burden of a 30-45-minute battery was reasonable. The goal of this and ongoing disease-specific studies is to develop a brief battery tailored to the study needs. Finally, participants also positively rated technical components of the app (e.g., screen size) and felt their data were private and secure. We also found a minimal impact of device on results, suggesting that the app is accessible across common smartphone types, an important factor for equitable implementation across demographic groups [35].

Prior to deploying cognitive tests to answer research or clinical questions, it is important to determine whether the tests produce similar results each time they are administered, a construct referred to as test-retest reliability. The design of this study, with three sessions conducted over two weeks, allowed for investigation of test-retest reliability. Results suggest the ALLFTD-mApp app cognitive tests demonstrated moderate to strong test-retest reliability, consistent with findings from a natural history study of FTD (ICC > 0.84) [11]. Similar levels of reliability have been observed in other unsupervised remote studies of healthy adults, such as the ARC app in cognitively normal older adults (ICC > 0.85 over 6-12 months; Nicosia et al., 2023), BRANCH in clinically normal mid- to late-life adults (r = 0.81; Papp et al., 2021), and the Neotiv app in healthy control, mild cognitive impairment and subjective cognitive decline cohorts (ICC = 0.80; Berron, Olsson, et al., 2024). Taken together, these results suggest that digital cognitive tests in healthy controls, even when administered in an unsupervised format, produce consistent results.

In addition to reliability, the construct validity of these tasks was supported by consistent and statistically significant associations with demographic characteristics, subjective cognitive complaints, and independent measures of cognitive function. Specifically, older age, an established predictor of processing speed, executive, functioning, and memory, was strongly associated with performance on mobile app tasks in this registry cohort, a result shown across other mobile testing platforms. Similar age-related trends have been observed in other mobile platforms; for example, Neotiv memory tasks have shown expected slowing and lower scores in older adults, consistent with known patterns of cognitive aging [36]. In addition, higher levels of education, another established correlate of cognitive performance, was associated with higher scores. Beyond these demographic factors, self- and informant-reported complaints are often one of the earliest indicators of cognitive changes and in some cases a bellwether for future cognitive decline [18]. In this study, overall subjective complaints, as well as domain-specific concerns in memory and executive functioning, were associated with app performance on measures of the same domain and across domains. Interestingly, associations were stronger with informant reports of cognition than self-reports, and self- and informant-scores were only modestly correlated, a previously reported phenomenon [37,38]. This may reflect differing perceptions, a lack of awareness of cognitive deficits in the participants, or could relate to methodological differences, as informants completed the 41-item ECog while participants completed the 12-item version. Notably, the two self-reported formats were highly correlated in our data. Finally, ALLFTD-mApp tasks were associated with performance on CBB tasks of the same domains and across domains. This bolsters recently reported results showing these mobile app tasks were associated with gold-standard neuropsychological tests that had been administered in person, as well as with clinician estimates of disease severity and brain volume [8,11,33,39,40]. Similar results of construct validity have been reported across several other unsupervised remote testing platforms [7,8,33,36,41,42].

### Limitations

Although overall recruitment was successful, younger adults, men, and individuals from racially and ethnically diverse backgrounds were underrepresented in the cohort. Culturally tailored materials were used to support outreach of underrepresented racial and ethnic groups, but these efforts did not substantially improve ethnic and racial representation. Future studies should prioritize improved representation to ensure that app-based cognitive assessments are applicable across different populations. Although this initial study focused on English-speaking participants, the app has recently been translated into several other languages and validation studies are underway, along with a study of app performance in participants from underrepresented racial/ethnic groups who endorse early-onset behavioral and cognitive concerns. In addition, the use of BHR to recruit participants may limit generalizability as participants are often highly motivated to join research and tend to be more technologically experienced than the broader population.

Testing in this study was not supervised, potentially introducing variability in testing environments. However, approximately 77% of participants completed each session without reporting distractions, similar to rates reported in other smartphone studies [7] and higher than rates observed in the FTD natural history cohort reported by Staffaroni et al. [11], reinforcing the feasibility of unsupervised testing and the reliability of data collected in real-world environments.

A final limitation is that the cognitive status of this registry-recruited cohort was defined primarily using subjective reports (i.e., cognitive complaints; self-reported diagnoses and medications). Prior work with the ALLFTD-mApp has defined cognitive status based on multidisciplinary consensus [11,12] but this was not available in the current work. Future studies should consider brief cognitive screeners (e.g., MoCA or Cogstate), validated semi-structured interviews for quantifying disease severity (e.g., electronic Clinical Dementia Rating [eCDR]; [43]), and biomarkers to improve participant characterization. Although we attempted to collect study partner ratings on all participants, the low response rate is a limitation of this study, although it is comparable to the BHR Study Partner Portal enrollment rate [37].

## Conclusion

The ALLFTD-mApp is a scalable, valid, and feasible tool for remote cognitive assessment. Its strong accuracy, reliability, and adherence—combined with meaningful associations with subjective cognitive concerns and independent cognitive tasks—support its potential for use in remote cognitive research. These findings advance the field of digital cognitive assessments and highlight a potential tool for remote monitoring of brain health in healthy and at-risk populations.

## Data Availability

Please contact authors regarding data availability

## Acknowledgements

This manuscript was prepared using a data set obtained from the UCSF Brain Health Registry (The Department of Radiology and Biomedical Engineering, The Regents of the University of California, Dr. Michael Weiner, Principal Investigator) and does not necessarily reflect the opinions or views of the UCSF Brain Health Registry investigators, the NIH or the private funding partners.

