## Supplemental Materials for "Remote, self-administered, smartphone cognitive testing in a registry-based cohort: Feasibility, reliability, and validity findings"

ALLFTD-mApp Composite Development

**METHODS:**

Participants were 1542 individuals from the associated manuscript as well as 378 additional participants from the ARTFL LEFFTDS Longitudinal Frontotemporal Lobar Degeneration (FTLD) consortium (ALLFTD). The ALLFTD sample includes participants with familial and sporadic FTLD spectrum disorders. They present with a heterogenous set of symptoms, suspected underlying pathology, clinical severity, and disease state (Heuer et al., 2023). The subsample of ALLFTD participants with smartphone data has been described previously (Staffaroni et al., 2024; Taylor et al., 2023). From a psychometric perspective, including ALLFTD participants and the BHR participants enables the factor to capture a broad range of the underlying latent cognitive ability.

Using structural equation modeling and item response theory, we created a measure of cognitive ability from four smartphone tasks: Color Clash (Stroop task of cognitive inhibition), Ducks in a Pond (Flanker task of attentional control), Animal Parade (N-back task of working memory), and Humi's Bistro (episodic memory). All participants completed at least one of these measures. First, all continuous data was recoded as ordinal scores with up to twelve response categories and at least five observations in each category (Mukherjee et al., 2020). Second, a scree plot was generated to inform model building decisions. Next, a confirmatory factor model was fit with a single general smartphone cognitive performance factor via item response theory using a graded response method (Samejima, 2011). Model fit was determined via factor loadings (>0.4 retained; >0.5 significant; Matsunaga, 2010; Williams et al., 2010), the Tucker Lewis Index (TLI; >0.90 acceptable, ≥ 0.95 excellent; Mukherjee et al., 2020; Schumacker & Lomax, 2004), the Comparative Fit Index (CFI; >0.90 acceptable, ≥ 0.95 excellent; Mukherjee et al., 2020; Savalei & Bentler, 2006), and Root Mean Square Error of Approximation (RMSEA; < 0.10 acceptable; MacCallum et al., 1996).

All analyses were completed in R (R Core Team & Team, 2023). Structural equation modeling and item response theory models were completed with the Lavaan (Rosseel, 2012) and MIRT (Chalmers, 2012) packages using a Maximum Likelihood (ML) estimator. Factor scores were generated for all participants using the "fscores" function within the MIRT package (estimation method = expected a-posteriori; quasi-Monte Carlo integration). This function produces a score for each participant.

**RESULTS**:

The exploratory factor analysis and associated scree plot suggested a one factor model best fit the data (Supplemental figure 1). As such, the confirmatory factor model included a single latent variable that loaded onto all observed smartphone tasks. This model demonstrated acceptable fit, as evidenced by acceptable fit across fit statistics (TLI=0.943; CFI=0.981; RMSEA=0.098). Factor loadings were all above the predetermined magnitude threshold (>0.4; supplemental figure 2). The factor was normally distributed. It demonstrated excellent 3-month test-retest reliability (r=0.88; n=153).

| | Overall | BHR | ALLFTD |
|---|---|---|---|
| **n** | 1936 | 1542 | 378 |
| **Age** | 53.60 (14.32) | 61.67 (7.35) | 53.30 (14.44) |
| **Years Education** | 16.32 (2.34) | 16.31 (2.33) | 16.33 (2.39) |
| **Race** | | | |
| *Caucasian* | 1711 (88.4) | 1351 (87.6) | 360 (95.2) |
| *Multiracial* | 79 (4.1) | 79 (5.1) | 0 (0.0) |
| *African American* | 45(2.3) | 41 (2.7) | 4 (1.1) |
| *Asian* | 52 (2.7) | 48 (3.1) | 4 (1.1) |
| *Native American* | 11 (0.6) | 9 (0.6) | 2 (0.5) |
| *Pacific Islander* | 1 (0.1) | 1 (0.1) | 0 (0.0) |
| *Other* | 13 (0.8) | 10 (0.7) | 3 (0.8) |
| *Unknown* | 24 (1.2) | 3 (0.2) | 5 (1.3) |
| **Gender (%)** | | | |
| *Male* | 621 (32.1) | 482 (31.3) | 139 (36.8) |
| *Female* | 1245 (64.3) | 1041 (67.5) | 204 (54.0) |
| *Other/Unknown* | 70 (3.6) | 19 (1.2) | 35 (9.3) |
| **CDR®+NACC-FTLD Global** | | | |
| *0* | 219 (11.3) | --- | 219 (57.9) |
| *0.5* | 69 (3.6) | --- | 69 (18.3) |
| *1+* | 79 (4.1) | --- | 79 (20.9) |
| *Unknown* | 1569 (81.0)* | --- | 11 (2.9) |

**Supplemental Table 1:** *Demographics of ALLFTD-mApp composite development subsamples*

Provides means (standard deviation) or count (percent) for demographics for the full sample (Overall), and each of the subsamples (BHR, ALLFTD) that were used to create the cognitive composite score.

* The CDR®+NACC-FTLD was not administered to the BHR sample.

**Acronyms**: **BHR**: Brain Health Registry; **ALLFTD**: ARTFL LEFFTDS Longitudinal Frontotemporal Lobar Degeneration (FTLD) consortium; **CDR®+NACC-FTLD Global:** Clinical Dementia Rating® Plus National Alzheimer's Coordinating Center Frontotemporal Lobar Degeneration Global Score.

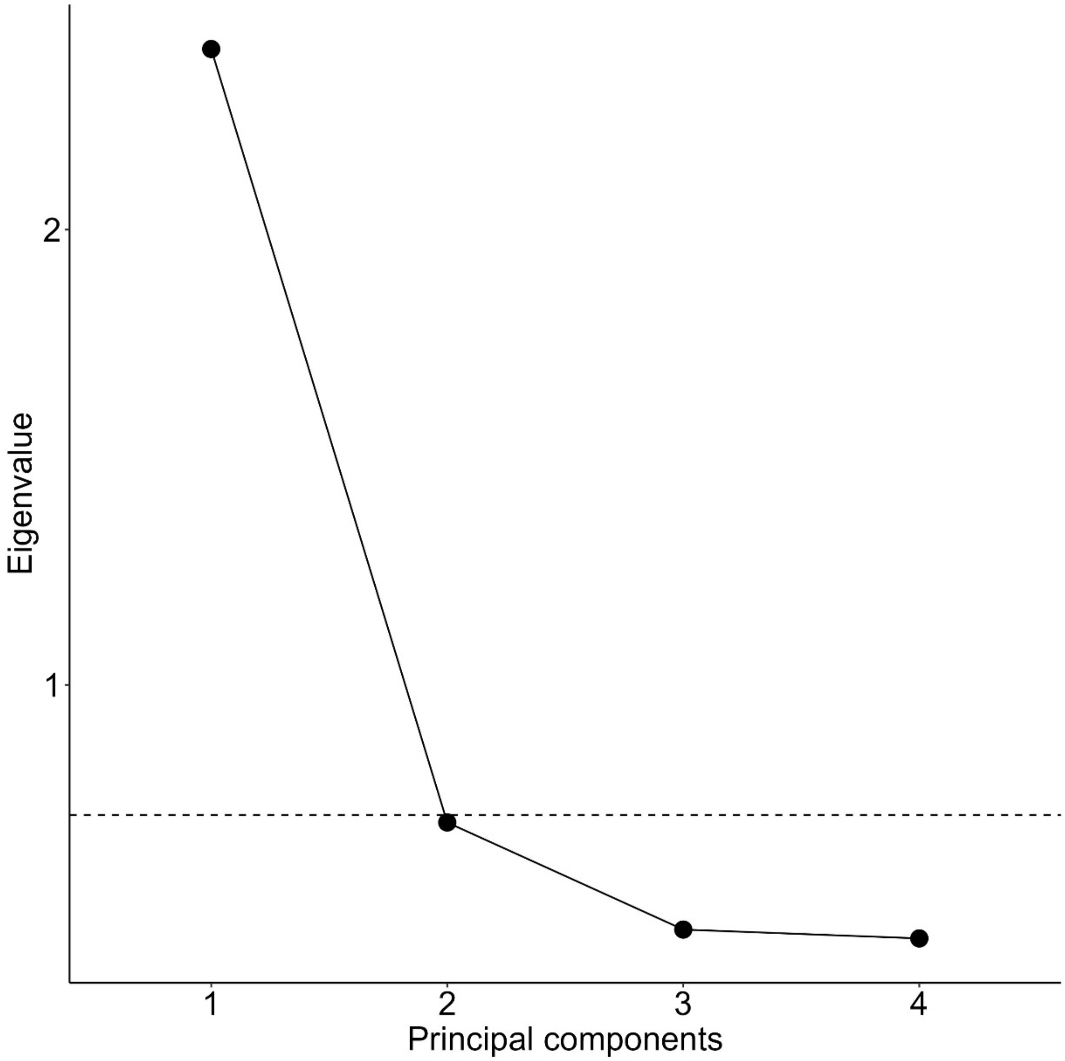


**Supplemental Figure 1**: *Scree plot*
This plot displays the number of principal components (X-axis) against the eigenvalues (Y-axis). A horizontal dashed line has been placed at the “elbow” of the scree plot to indicate the location of a sharp decline in eigenvalues.

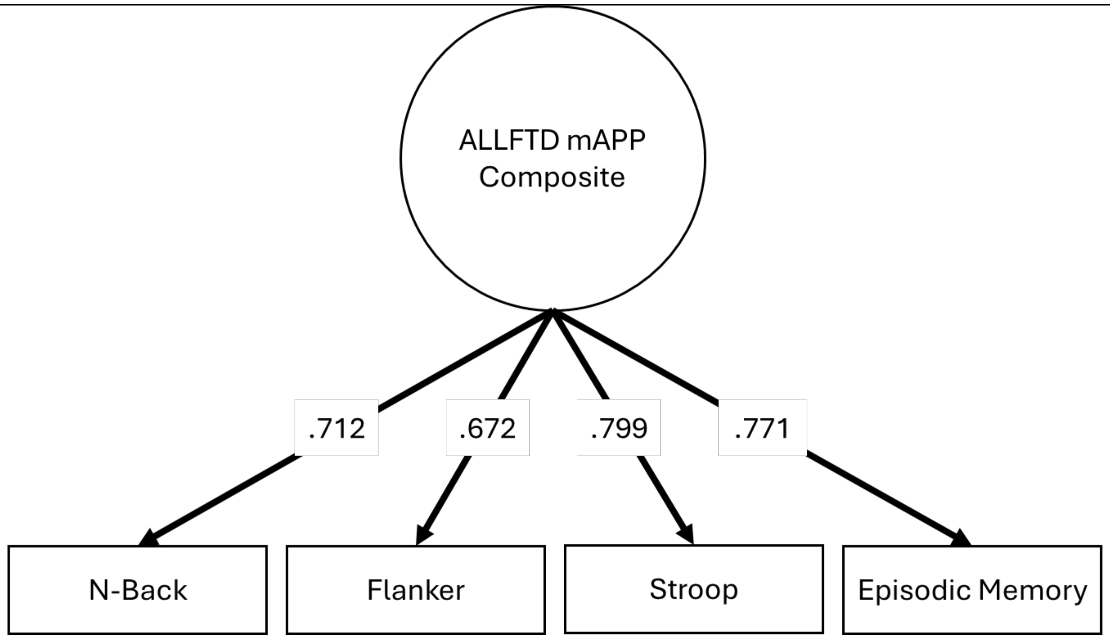


**Supplemental Figure 2**: *Confirmatory factor model*
This model presents unstandardized loadings between each ALLFTD mApp task (rectangle) and the latent factor (circle).

| | | Age Bin | | | | | | |
|---|---|---|---|---|---|---|---|---|
| | **Overall** | **18-29** | **30-39** | **40-49** | **50-59** | **60-69** | **70-79** | **80+** |
| **n** | 314 | 1 | 12 | 28 | 31 | 139 | 88 | 15 |
| **Age (mean, SD)** | 64.58 (11.00) | 24.88 (NA) | 36.95 (2.62) | 45.70 (2.75) | 56.18 (2.47) | 65.28 (2.79) | 73.28 (2.72) | 84.35 (3.59) |
| **Years Education** | 16.42 (2.38) | 16.00 (NA) | 17.33 (1.56) | 16.79 (1.97) | 17.13 (1.89) | 16.08 (2.40) | 16.42 (2.66) | 16.80 (2.21) |
| **Race (%)** | | | | | | | | |
| *Caucasian* | 269 (85.7) | 0 (0.0) | 9 (75.0) | 19 (67.9) | 27 (87.1) | 121 (87.1) | 78 (88.6) | 15 (100.0) |
| *Multiracial* | 33 (10.5) | 0 (0.0) | 3 (25.0) | 7 (25.0) | 3 (9.7) | 12 (8.6) | 8 (9.1) | 0 (0.0) |
| *African American* | 3 (1.0) | 0 (0.0) | 0 (0.0) | 0 (0.0) | 0 (0.0) | 2 (1.4) | 1 (1.1) | 0 (0.0) |
| *Asian* | 6 (1.9) | 1 (100.0) | 0 (0.0) | 2 (7.1) | 0 (0.0) | 2 (1.4) | 1 (1.1) | 0 (0.0) |
| *Other* | 2 (0.6) | 0 (0.0) | 0 (0.0) | 0 (0.0) | 0 (0.0) | 2 (1.4) | 0 (0.0) | 0 (0.0) |
| *Pacific Islander* | 1 (0.3) | 0 (0.0) | 0 (0.0) | 0 (0.0) | 0 (0.0) | 0 (0.0) | 0 (0.0) | 0 (0.0) |
| **Female Gender (%)** | 188 (59.9) | 0 (0.0) | 11 (91.7) | 20 (71.4) | 20 (64.5) | 84 (60.4) | 46 (52.3) | 7 (46.7) |
| **Cognition** | | | | | | | | |
| *Study Partner ECog-41* | 1.29 (0.45) | NA | 1.07 (0.12) | 1.20 (0.22) | 1.14 (0.16) | 1.36 (0.57) | 1.30 (0.37) | 1.27 (0.29) |

**Supplemental Table 2:** *Study partner subsample*

This table presents the demographics of the subset of participants whose study partners completed the ECog-41 questionnaire (N = 314). The average informant–reported ECog-41 score was 1.29 (SD = 0.45), indicating generally low levels of reported functional difficulties. This subsample was used in secondary analyses evaluating the relationship between participant characteristics and study partner–reported cognitive concerns.

| Task | Session Comparison | $\rho$ | 95% CI | p |
|---|---|---|---|---|
| Stroop | 1–2 | 0.78 | [0.75, 0.80] | < 0.001 |
| | 2–3 | 0.85 | [0.84, 0.87] | < 0.001 |
| Flanker | 1–2 | 0.82 | [0.79, 0.84] | < 0.001 |
| | 2–3 | 0.84 | [0.81, 0.86] | < 0.001 |
| Episodic Memory | 1–2 | 0.67 | [0.64, 0.70] | < 0.001 |
| | 2–3 | 0.70 | [0.67, 0.74] | < 0.001 |
| N-Back | 1–2 | 0.67 | [0.64, 0.71] | < 0.001 |
| | 2–3 | 0.71 | [0.68, 0.74] | < 0.001 |
| Composite | 1–2 | 0.80 | [0.78, 0.82] | < 0.001 |
| | 2–3 | 0.85 | [0.83, 0.87] | < 0.001 |

**Supplemental Table 3:** *Test-retest reliability of ALLFTD-mApp cognitive tasks across sessions*

This table presents the Spearman's correlations ($\rho$) with 95% confidence intervals are reported for each cognitive task and overall composite, comparing performance between Session 1-2 and Session 2-3. All correlations were statistically significant at $p < .001$, indicating moderate to strong reliability across tasks.
